# Plant-based natural products for symptomatic relief of Parkinson’s disease: prevalence, interest, awareness and determinants

**DOI:** 10.1101/2023.11.17.23298666

**Authors:** Sandra Diadhiou, Bart R. Maas, Sabine Schootemeijer, Bastiaan R. Bloem, Nienke M. de Vries, Frédéric Calon, Sirwan K.L. Darweesh, Aurelie de Rus Jacquet

## Abstract

Natural health products (NHP) have emerged as a potential symptomatic therapeutic approach for persons with Parkinson’s disease (PwP). The objective of this study was to quantify the prevalence of ever use of NHP, interest in plant-based NHP, awareness of potential herb-drug interactions, and how often NHP use was discussed by PwP with their healthcare professionals. We addressed these objectives by embedding a cross-sectional 4-item survey within a large population-based cohort of PwP (PRIME-NL study). Sixty-five percent (n=367) of the 566 participants who were contacted completed the survey. Of those participants, 132 (36%) reported having used NHP to alleviate Parkinson’s disease (PD)-related symptoms, with coffee, cannabis and turmeric being the most popular. Overall, 12% (n=44) of PwP had used at least one NHP other than coffee or cannabis. Furthermore, 71% (n=259) participants expressed an interest in exploring the use of NHP, but only 39% (n=51) of NHP users were aware that these products could interact with PD medication. Finally, only 39% (n=51) of NHP users had discussed the use of NHP with their neurologist or PD nurse specialist. In a sensitivity analysis, we conservatively assumed that all non-responders to the survey had never used NHP and had no interest in exploring NHP. This rendered an estimated prevalence of NHP use of 23% and an estimated interest in exploring NHP of 46%. In conclusion, over one in three PwP has used NHP to alleviate symptoms of PD and the majority of PwP is interested in exploring the use of plant-based NHP. Most users had not discussed the intake of NHP with their PD healthcare professional and were unaware that these products could interact with PD medication. This study supports the need for evidence-based research on the properties of plant-derived therapeutics.

---

Parkinson’s disease (PD) is the world’s fastest growing neurodegenerative disorder, currently affecting more than 8.5 million people worldwide ^1, 2^. Life-disrupting clinical symptoms include motor symptoms (e.g. loss of balance, gait impairment, tremor) as well as non-motor symptoms (e.g. anxiety, constipation, cognitive decline) ^4, 5^. Available therapeutic approaches are limited to symptomatic treatments, such as those compensating for dopamine deficits induced by the loss of dopaminergic neurons^3^. The limited therapeutic options available to relieve clinical symptoms, the known side effects of PD medication, and the increasing attention for non-pharmacological interventions may encourage people with PD (PwP) to explore the use of plant-based supplements to mitigate their symptoms^4-8^. A number of natural health products (NHP) are commercially available. Their therapeutic indications often derive from ancestral knowledge documented in local pharmacopoeia, although regulations and classifications vary according to the country of commercialization. Indeed, the practice of traditional herbal medicine can be a primary source of healthcare for patients across Asia and Africa ^9–11^. In the European Union (EU), three main regulatory pathways are offered to market herbal medicinal products, including Traditional Use Registration, Well-Established Use Marketing Authorization, and Stand-Alone or Mixed Applications ^9^. These regulatory pathways impose varying requirements depending on the existence of established traditional uses within the EU, and/or scientific evidence of safety and efficacy.

There is a lack of systematic studies in a population-representative sample of PwP that quantified the prevalence of ever use of NHP to mitigate PD-related symptoms, explored the interest in using NHP or the determinants of this interest, and evaluated the awareness of potential herb-drug interactions. The objective of this cross-sectional study in a representative sample of the spectrum of PwP was to address these gaps in knowledge. Specifically, we evaluated the proportion of PwP who had used or are interested in exploring plant-based natural products to mitigate PD-related symptoms. We also quantified the awareness of potential herb-drug interactions among NHP users, and the proportion of PwP who had discussed the use of plant-based products with their neurologist or PD nurse specialist. A secondary objective was to identify determinants of interest in NHP, including gender identity, age, education level, disease duration, and Hoehn and Yahr (H&Y) stage.

## Methods

### Participants

This cross-sectional survey was embedded in the Proactive and Integrated Management and Empowerment of Parkinson’s Disease – Netherlands (PRIME-NL) study ^10^. The PRIME-NL study is an ongoing observational cohort study that evaluates the utility of a new integrated care model (PRIME Parkinson care) using two data sources: pseudoanonymized medical claims data on the complete population and annual questionnaires. The current survey is embedded in the questionnaire subcohort. The questionnaires are administered annually, including a baseline assessment (in 2020) and five annual follow up-assessments (2021-2025). The PRIME-NL study has been approved by the Ethical Board of the Radboud University Medical Center. Participants in the PRIME-NL study were recruited through ParkinsonNEXT (an online platform to connect PwP and researchers in the Netherlands)^11^, the Dutch Parkinson patient association^12^, and neurologists in the PRIME Parkinson care region. All participants provided digital or written informed consent before inclusion in the study. The only inclusion criterion was having a clinical diagnosis of parkinsonism. Diagnosis of all participants was established by a neurologist. Exclusion criteria at the time of enrolment were: parkinsonism caused by medication, receiving treatment at a university medical center, or not having visited the neurology outpatient clinic in the year prior to inclusion. In the PRIME-NL cohort, 920 of 984 participants had PD, while 64 participants had a form of atypical parkinsonism. For this report, we only included PwP and excluded people with atypical parkinsonism. A previous study showed that the characteristics of the PRIME-NL cohort are largely representative of the full spectrum of PwP in the Netherlands, with the exception of a slightly lower age (69.6 years compared to 72.7 years) and a slightly longer disease duration (6.2 years compared to 5.3 years)^13^.

### Questionnaire

The 566 participants who responded to the PRIME-NL questionnaires between July 2022 and May 2023, i.e. in the second and third follow-up years, were offered to answer the NHP survey. Questionnaires were self-administered by the participants, either electronically (n=307), on paper (n=58) or by a well-trained research employee over the phone (n=2). The 4-item custom questionnaire was administered to document the use of plant-based natural products by PwP to specifically attenuate PD-related symptoms (full questionnaire available in **Supplementary File 1**). The questions were designed to (1) establish the prevalence of usage of herbal remedies among PwP, (2) gauge their interest in plant-based natural products, (3) assess whether PwP communicate with their PD specialist regarding NHP, and (4) evaluate their awareness of potential herb-drug interactions. A total of 11 herbal supplements were included in the study, based on their availability to PwP living in The Netherlands and their potential benefits to mitigate motors and non-motor symptoms (see **Supplementary Information** for details of the potential benefits of these NHP). An option “other” was provided to record the use of additional NHP. The study did not discriminate between food and supplements. Question items regarding the intake of herbal supplements were phrased to ensure that participants only reported those used to alleviate PD-related symptoms, and not recreational or non-medical uses. Ascertainment methods of covariates have previously been published in detail ^10^. In short, gender identity of participants was assessed by offering the options “men”, “women”, and “other”, and none of the respondents selected the “other” option. Hoehn and Yahr stages were based on items in self-administered questionnaires. Education level was self-administered and recategorized into three categories (low-, medium-, and high education) according to the International Standard Classification of Education^14^.

### Statistical analyses

We quantified the prevalence of ever use of NHP, interest and determinants of NHP use, awareness of potential herb-drug interactions, and the prevalence of discussing NHP use with PD healthcare professionals among PwP using descriptive statistics. Categorical data were analyzed using a Pearson exact Chi-square test with Monte Carlo estimation when appropriate. Continuous data were analyzed using a Student’s t-test. Descriptive statistics were performed using the SPSS software (IBM SPSS Statistics for Windows version 27.0, IBM Corp., Armonk, NY, USA) with the significance level set at p ≤ 0.05. We also performed several sensitivity analyses. First, we performed a sensitivity analysis to evaluate the proportion of NHP users and the percentage of participants interested in learning more about these products, in which we conservatively assumed that none of the non-responders had ever used or had any interest in exploring plant-based treatments. Second, we determined the external validity of the sample of participants by comparing both demographical and clinical characteristics of responders versus non-responders. Third, we performed a sensitivity analysis to evaluate the proportion of participants who used NHP other than coffee and cannabis. Fourth, we determined how many participants used multiple products, as illustrated in Venn diagrams and a frequency histogram. Additional analyses were performed to identify determinants of NHP use, and consisted in a multivariate Logistic Regression model with consumption of herbal remedies as dependent variable and gender identity, age, education level, disease duration, and Hoehn and Yahr stage as independent variables. We determined β-coefficients and p-values for the independent variables and considered p<0.05 as significant association. Regression analysis was performed in R Statistics.

## Results

### Demographics

We invited 566 participants and 367 (65%) responded to the questionnaire. The cohort of participants (n=367) vs. non-participants (n=199) was comparable in terms of age, education level and stratification by Hoehn and Yahr stages, but the group of participants consisted of more women vs. men (p=0.005) and an overall shorter disease duration (p<0.001) (**Supplementary Table 1**). Demographic evaluation of the cohort of participants revealed an average 70.3 ± 8.1 years of age and 7.1 ± 4 years of disease duration. A total of 72% (n=166) of women contacted responded to this survey, compared to 60% (n=201) of men. Women participants were younger (p=0.046), had a longer disease duration (p<0.001) and greater average Hoehn & Yahr scores (p=0.039) (**Supplementary Table 2**). Men and women had comparable level of medium-level education (men (%), 23; women (%), 27), but a greater proportion of men reported a higher-level education (men (%), 58; women (%), 45) (**Supplementary Table 2**).

### Prevalence of NHP consumption

Overall, 36% of participants reported having ever consumed at least one of the 11 herbs listed in the survey or additional herbal products listed as “other”. Sixteen percent of participants used coffee specifically for medicinal purpose, 13% used cannabis, and 5% reported using both coffee and cannabis products. Other herbal supplements used by PwP included turmeric (*Curcuma longa*; 10%), chamomile (*Matricaria* sp., *Chamaemelum* sp.; 5%), and velvet bean (*Mucuna pruriens*; 7%) (**Table 1**). All other supplements were reported by less than 2% of participants. These 11 NHP were used as single product by 22% (n=80) PwP, 13% (n=46) ever used more than one NHP, and 1% (n=4) reported using up to 5 NHP (**Figure 1, Supplementary Figure 1**). A sensitivity analysis showed that, overall, 12% (n=44) of participants consumed NHP other than cannabis products or coffee, and the most commonly used NHP by this subgroup were turmeric (n=20), velvet bean (n=13) and chamomile (n=8) (**Table 1**). Sensitivity analyses were performed to estimate the use of NHP in the entire cohort of participants (n=367) and non-participants (n=199). Assuming that the entire subgroup of non-responders did not use herbal products, we estimate that, in this cohort, 23% of PwP (132/566) used NHP to mitigate PD-related symptoms.

**Table 1.** Use of plant-based supplements and natural products to treat symptoms associated with PD. The p-values were calculated to compare the responses by gender identity. Abbreviation: n, number. ^a^In the sensitivity analysis, participants who consumed cannabis, coffee, or both, were excluded and data was not stratified by gender.

| Have you ever used any of the following herbal remedies with the purpose of alleviating symptoms related to Parkinson’s disease? |  |  |  |  |  |
| --- | --- | --- | --- | --- | --- |
| Proportion of participants who consumed at least one of the herbs listed below, % (n) | 36 (132) |  |  |  |  |
|  | Total (n=367) | Men (n=201) | Women (n=166) | P-value | Sensitivity<sup>a</sup> (n=44) |
| Cannabis. Yes % (n) | 13.4 (48) | 10.4 (21) | 16.3 (27) | 0.100 | - |
| Coffee. Yes % (n) | 16 (58) | 13.9 (28) | 18.1 (30) | 0.279 | - |
| Both cannabis and coffee. Yes % (n) | 5.1 (18) | 3.0 (6) | 7.2 (12) | 0.061 | - |
| Chamomile. Yes % (n) | 4.8 (17) | 3.0 (6) | 6.6 (11) | 0.099 | 8 |
| Curcumin. Yes % (n) | 1.1 (4) | 0.5 (1) | 1.8 (3) | 0.229 | 1 |
| Guarana. Yes % (n) | 0 | 0 | 0 | - | 0 |
| Passion flower. Yes, % (n) | 1.1 (4) | 1.0 (2) | 1.2 (2) | 0.847 | 1 |
| Resveratrol. Yes, % (n) | 0.9 (3) | 0.5 (1) | 1.2 (2) | 0.454 | 1 |
| Rhodiola. Yes, % (n) | 1.1 (4) | 0.5 (1) | 1.8 (3) | 0.229 | 1 |
| St John’s Wort. Yes, % (n) | 1.1 (4) | 1.5 (3) | 0.6 (1) | 0.414 | 1 |
| Tumeric. Yes, % (n) | 9.6 (35) | 9.5 (19) | 9.6 (16) | 0.952 | 20 |
| Velvet bean. Yes, % (n) | 6.9 (25) | 6.0 (12) | 7.8 (13) | 0.481 | 13 |
| Other. Yes, % (n) | 5.9 (21) | 4.5 (9) | 7.2 (12) | 0.259 | 10 |
| Would you be interested in learning more about possible herbal remedies to alleviate certain symptoms related to Parkinson’s disease? |  |  |  |  |  |
|  | Total (n=367) | Men (n=201) | Women (n=166) | P-value |  |
| Across the entire cohort, Yes, % (n) | 70.6 (259) | 70.1 (141) | 71.1 (118) | 0.845 |  |
| Have you ever discussed the possible use of herbal remedies with your neurologist or Parkinson’s disease nurse specialist? |  |  |  |  |  |
|  | Total (n=367) | Men (n=201) | Women (n=166) | P-value |  |
| Among participants who declared not using herbs. Yes % (n) | 7.0 (16) | 5.9 (8) | 8.0 (8) | 0.533 |  |
| Among participants who declared already using herbs. Yes % (n) | 38.7 (51) | 36.4 (24) | 40.9 (27) | 0.592 |  |
| Among participants who declared using velvet bean. Yes % (n) | 76.0 (19) | 75.0 (9) | 76.9 (10) | 0.910 |  |
| Are you aware that some herbal remedies and prescribed Parkinson’s disease medications may work together (or against each other)? |  |  |  |  |  |
|  | Total (n=132) | Men (n=66) | Women (n=66) | P-value |  |
| Among participants who declared already using herbs. Yes % (n) | 38.7 (51) | 30.3 (20) | 47.0 (31) | 0.049 |  |

**Figure 1.**
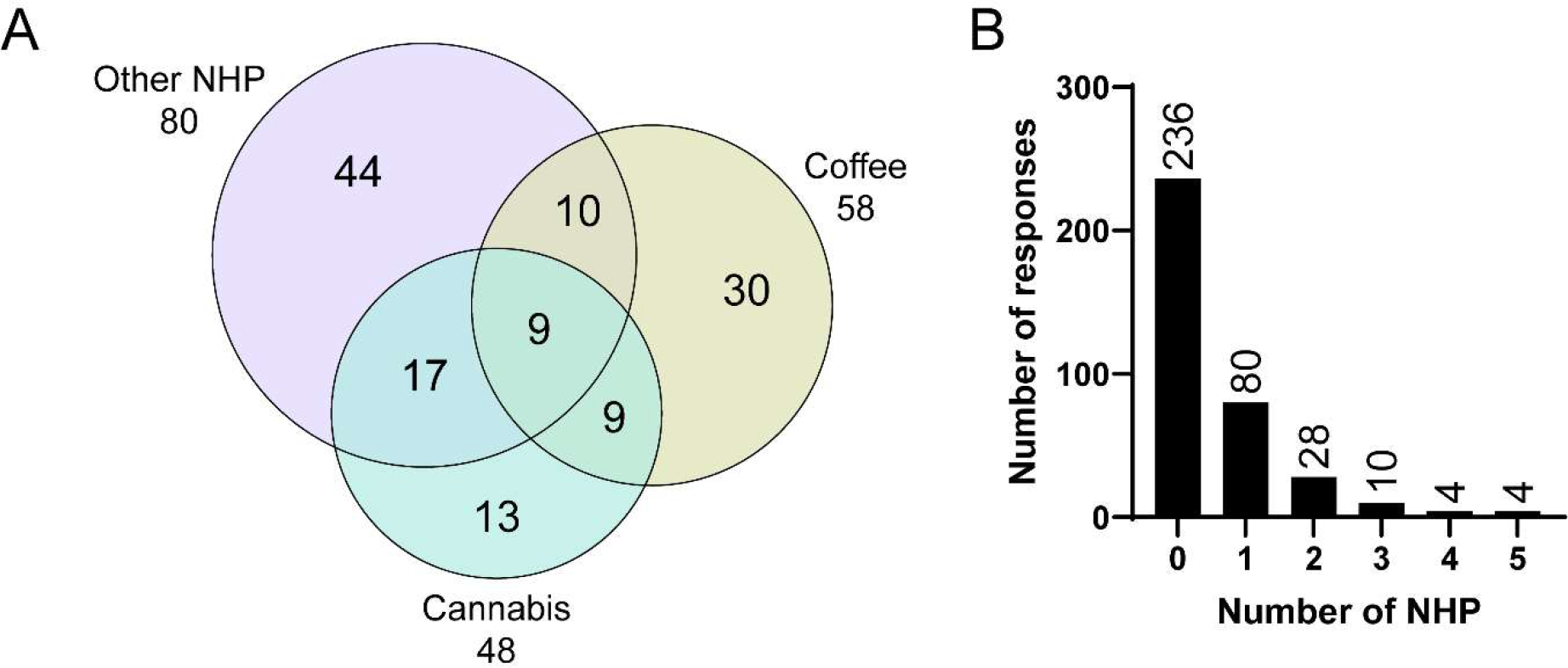
Overview of the usage of single vs. combination of NHP by PwP. **(A)** Venn diagram illustrating the number of participants who acknowledged using NHP listed in Table 1. Each circle represents the use of coffee, cannabis, or all other NHP combined. The numbers in these circles represent the number of participants using the corresponding NHP combination, while the total number of participants who use each NHP is provided outside the circles. **(B)** Histogram representing the frequency of NHP use. The “other” category is excluded from analysis. Abbreviation: NHP, natural health products.

### Expressed interest and determinants of NHP consumption

A total of 71% (n=259) of participants reported an interest to learn more about the use of plant-based supplements to treat PD-related symptoms. We performed a sensitivity analysis in which we assumed that the entire subgroup of non-responders was not interested in receiving additional information about NHP. The results show that 46% of PwP (259/566) in our cohort declared being interested in learning more about the use of NHP to mitigate PD-related symptoms. The consumption of herbal remedies in the cohort of participants was not significantly associated with gender identity (β=0.26, p=0.76), nor with age (β=-0.06, p=0.23), education level (β=-0.08, p=0.79), disease duration (β=-0.04, p=0.74) or H&Y stage (β=0.35, p=0.34).

### Awareness of potential NHP-drug interactions

When asked if participants were aware that some herbal remedies and prescribed PD medications could interact, 39% (n=51) of participants who declared using NHP acknowledged being aware of potential interferences, with a significantly greater proportion of women vs. men (p=0.049) (**Table 1**). Our study revealed that 39% of participants who declared using NHP had prior discussions with their neurologist or PD nurse specialist regarding this self-medication. Furthermore, velvet bean naturally contains L-DOPA (from 3 to more than 100 mg/dose depending on the source^15^) and could therefore interact with dopamine replacement therapies. Among participants who declared using velvet bean, 76% indicated that they had discussed the use of herbal remedies with their PD specialists.

## Discussion

This survey revealed that over one in three PwP had used NHP to alleviate symptoms of PD and the majority of PwP was interested in exploring the use of NHP. Most NHP consumers had not discussed the use of NHP with their PD healthcare professional (with the exception of those using velvet bean) and most were unaware that these products could interact with PD medication. This study supports the need for rigorous evidence-based research on the properties of plant-derived therapeutics, and also identifies a need for designing communication strategies to better inform PwP.

In this report, we documented that coffee, cannabis, and turmeric are popular plant-based products. The 13% of participants who reported having ever used cannabis to alleviate PD-related symptoms is comparable to a study in Norway, where 11% of PwP reported cannabis consumption ^16^, but lower compared to a survey conducted in the USA where 25% of PwP reported cannabis use across 49 states ^17^. Our analysis of external validity suggests that findings from the 367 PwP who agreed to take part in this survey could likely be generalized to other cohorts of similar age, education, and Hoehn & Yahr stage distribution, and we estimate the prevalence of ever use of NHP to be over one in five and the prevalence of interest over two in five PwP. Furthermore, 13% (n=46) of PwP ever used more than one NHP. Additional studies are needed to determine if these products are consumed concomitantly and may result in herb-drug interactions.

The interest in natural sources of bioactive molecules documented here corroborates other studies indicating that PwP seek to become active players in the design of their therapies ^18^. This offers an opportunity to further develop personalized treatments and promote patient involvement in health care decision making. However, it is important to caution patients against the use of potentially ineffective therapies that may delay the prescription of evidence-based treatments. For example, a case report detailed the self-use of velvet bean that caused avoidable disabilities by postponing the intake of a more effective treatment consisting of velvet bean combined with a dopa-decarboxylase inhibitor ^19^. Our findings also support the need for in-depth investigation of herb-drug interactions, and the symptomatic and disease-modifying properties of natural products. Natural supplements are available over the counter and do not require medical prescriptions, which gives an apparent impression of innocuity despite containing bioactive molecules. In this context, we found that only a subset of participants was informed about herb-drug interactions. These could, for example, affect the efficiency of PD medication via pharmacokinetic or pharmacogenomic interactions ^20, 21^. While more research is needed to characterize in detail how NHP may interact with commonly prescribed PD medication, cautionary measures advocate for the design of educational material to inform consumers that NHP available over the counter might potentially interact with prescribed pharmaceuticals. In particular, patients should be encouraged to openly discuss the use of NHP or any other supplements with their physician, so that a joint decision can be made about possible benefits and safety. Furthermore, the bioactive components of NHP may vary depending on agricultural and ecological variables, and this challenge will need to be addressed to further establish their clinical benefits. Additional studies are also needed to extend the survey to other countries/cultures, and identify which additional supplements are used by PwP (e.g. omega-3 polyunsaturated fatty acids ^22^).

The results discussed here present a number of limitations. The optional nature of the survey resulted in a 65% response rate, and these participants may have been more interested in the use of NHP compared to the general PwP population. Also, the short 4-item questionnaire does not allow for a thorough description of NHP practices such as frequency of use, NHP presentation (i.e. pills, tea), which PD-related symptoms are targeted, or impact of living environment on NHP consumption. Additional information on the proportion of PwP who use NHP as a replacement vs. in addition to PD medication is also needed to portray a complete profile of NHP use. These elements could be investigated further in follow-up studies.

## Conclusions

We report that many PwP in our sample seek herbal remedies to complement their prescribed PD medication, and most participants demonstrated an interest in learning about these products. However, there is a need for educational resources that educate PwP on the potential interactions between NHP and PD medication. We did not identify relevant determinants of NHP use.

## Data Availability

All data produced in the present study are available upon reasonable request to the authors

## Acknowledgements

The authors would like to thank all participants who completed this survey, and the PRIME-NL team involved in the project.

## Disclosures

### Financial Disclosures for the previous 12 months

A.d.R.J. is supported by a Launch award from the Parkinson’s Foundation (PF-Launch-937655), the Michael J Fox Foundation, internal funds from the Institute of Nutrition and Functional Foods (INAF), the Quebec Parkinson’s Network, and funds from the Fondation CHU de Québec. B.R.B. currently serves as Editor in Chief for Journal of Parkinson’s disease; serves on the editorial board of Practical Neurology and Digital Biomarkers; has received honoraria from serving on the scientific advisory board for AbbVie, Biogen, and UCB; has received fees for speaking at conferences from AbbVie, Zambon, Roche, GE Healthcare, and Bial; and has received research support from the Netherlands Organization for Scientific Research, The Michael J. Fox Foundation, UCB, AbbVie, the Stichting Parkinson Fonds, the Hersenstichting Nederland, the Parkinson’s Foundation, Verily Life Sciences, Horizon 2020, the Topsector Life Sciences and Health, the Gatsby Foundation, and the Parkinson Vereniging. N.M.d.V reports grants from The Netherlands Organisation for Health Research and Development (ZonMw) and The Michael J Fox Foundation. F.C. is a Fonds de recherche du Québec-Santé research scholar and is supported by the Canadian Institutes of Health Research. S.K.L.D. has received funding from the Parkinson’s Foundation (PF-FBS-2026), ZonMW (09150162010183), ParkinsonNL (P2022-07 and P2021-14), Michael J Fox Foundation (MJFF-022767) and Edmond J Safra Foundation.

