## Supplementary Information for "Plant-based natural products for symptomatic relief of Parkinson’s disease: prevalence, interest, awareness and determinants"

### Plant-based natural products to treat Parkinson's disease-related symptoms: prevalence, interest, awareness and determinants

Sandra Diadhiou[1,2,3, #], Bart R. Maas[4, #], Sabine Schootemeijer[4], Bastiaan R. Bloem[4], Nienke M. de Vries[4], Frédéric Calon[1,2,3], Sirwan K.L. Darweesh[4], Aurelie de Rus Jacquet[2,3,5]

[1] Faculty of Pharmacy, Université Laval, Canada

[2] Axe neurosciences, centre de recherche du CHU de Québec-Université Laval, Canada

[3] Institute of Nutrition and Functional Foods, Université Laval, Canada

[4] Radboud University Medical Center; Donders Institute for Brain, Cognition and Behavior; Department of Neurology; Center of Expertise for Parkinson & Movement Disorders; Nijmegen, The Netherlands

[5] Department of Psychiatry and Neurosciences, Faculty of Medicine, Université Laval, Canada

[#] Contributed equally

Correspondence:
Aurelie de Rus Jacquet, PhD


**Discussion on the benefits of natural health products (NHP) to mitigate motor and non-motor symptoms experienced by people with Parkinson's disease (PwP)**

The results from the study presented herein can inform on how NHP may serve PwP improve their quality of life. We included the following 11 NHP based on their potential to mitigate PD-related motor and non-motor symptoms, and the survey did not distinguish between supplements and foods.

*Cannabis products*. One of the products of interest is medical cannabis, for which randomized and non-randomized clinical trials have been conducted. The results suggest a potential attenuation of the severity of motor (e.g. tremor) and non-motor (e.g. sleep, pain) symptoms over the course of several weeks of treatment [1].

*Chamomile*. Chamomile is traditionally used in the treatment of mild anxiety and sleep disorders [2-5], and could therefore attenuate non-motor symptoms experienced by PwP.

*Coffee*. We inquired about coffee consumption, which has been associated with a reduced risk of developing PD and symptom reduction [6, 7]. Studies suggest that caffeine is the bioactive molecule largely responsible for these effects, as drinking decaffeinated coffee abrogates the benefits, and other caffeine-containing products also attenuate motor and non-motor symptoms[7-9].

*Turmeric*. This Indian spice is enriched in curcumin, a polyphenolic compound largely studied for its beneficial effects on brain health [10], and under extensive investigation to translate promising *in vitro* and *in vivo* neuroprotective activities to the clinic [11, 12].

*Velvet bean*. Velvet bean has been explored as a natural source of L-DOPA, with studies estimating concentrations ranging from 4 to 7 % of the dried seeds [13, 14]. Clinical use of velvet bean to replace L-DOPA therapy has been reported to induce beneficial effects on motor symptoms [15-17].

*Other NHP of interest*. An additional three NHP were included in this survey for their traditional uses as psychoactive (guarana, *Paullinia cupana*, an alternative source of caffeine), calming (passionflower, *Passiflora incarnata*), or antidepressant (St John's Wort, *Hypericum perforatum*; rhodiola, *Rhodiola rosea*) agents [18-22], and might therefore mitigate non-motor PD symptoms.

The study revealed that respondents more often reported crude herbal preparations (e.g. turmeric) than isolated natural products found in these plants (e.g. curcumin) (**Table 1**). Published reports suggest that isolated natural compounds, in particular curcumin, are less bioavailable compared to the crude preparation[23, 24]. It is unclear why turmeric was favored compared to curcumin, but a possibility is PwP's prior knowledge of these physiological differences or the preference for food products vs. commercial supplements. Overall, NHP are bioactive molecules that could alter patients' responses to a therapeutic treatment, and a significant concern is the concomitant use and potential interference between PD medication and herbal supplements, which could lead to decreased potency of the prescribed treatment and/or detrimental herb-drug interactions. Future studies could investigate the preferential use of food vs. supplements by PwP, and evaluate the prevalence of NHP-drug interactions in patient populations.

**Supplementary Table 1. Comparison of characteristics between participants and non-participants to the survey**.

| | Participants (n=367) | Non-participants (n=199) | P-value |
|---|---|---|---|
| **Demographics** | | | |
| Gender identity, men, % (n) | 54.8 (201) | 66.8 (133) | 0.005 |
| Age, years, mean (SD) | 70.3 (8.1) | 70.7 (7.9) | 0.582 |
| **Education, % (n)** | | | |
| Low | 22.6 (83) | 27.1 (54) | 0.527 |
| Medium | 24.8 (91) | 25.6 (51) | |
| Higher | 52.0 (191) | 46.2 (92) | |
| **Disease severity** | | | |
| Disease duration, years, median (IQR) | 6.2 (4.6) | 7.2 (5.5) | <0.001 |
| H&Y (1-5 scale), mean (SD) | 2.5 (1.1) | 2.55 (1) | 0.600 |
| H&Y=1, % (n) | 22.6 (81) | 15.0 (29) | 0.164 |
| H&Y=2, % (n) | 34.1 (122) | 38.9 (75) | |
| H&Y=3, % (n) | 21.8 (78) | 23.3 (45) | |
| H&Y=4, % (n) | 18.4 (66) | 21.2 (41) | |
| H&Y=5, % (n) | 3.1 (11) | 1.6 (3) | |

Comparison of the demographics and clinical characteristics of responders (PRIME-NL participants who completed the survey) and non-responders (PRIME-NL participants who did not agree to respond to the survey). Mean and standard deviation are reported, unless noted otherwise. Abbreviation: H&Y, Hoehn and Yahr; IQR, interquartile range; SD, standard deviation; n, number.

**Supplementary Table 2. Patient demographics**.

| **Demographics** | | | | |
|---|---|---|---|---|
| Gender identity, men, % (n) | 54.8 (201) | | | |
| | Total (n=367) | Men (n=201) | Women (n=166) | P-value |
| Age, years, mean (SD) | 70.3 (8.1) | 71.1 (7.9) | 69.4 (8.3) | 0.046 |
| **Education, % (n)** | | | | |
| | Total (n=367) | Men (n=201) | Women (n=166) | P-value |
| Low | 23.0 (83) | 18.9 (38) | 27.1 (45) | 0.089 |
| Medium | 25.0 (91) | 22.9 (46) | 27.1 (45) | |
| Higher | 51.5 (191) | 57.7 (116) | 45.2 (75) | |
| **Disease severity** | | | | |
| | Total (n=367) | Men (n=201) | Women (n=166) | P-value |
| Disease duration, years, median (IQR) | 6.2 (4.6) | 5.6 (3.7) | 6.9 (5.7) | <0.001 |
| H&Y (1-5 scale), mean (SD) | 2.5 (1.2) | 2.3 (1.1) | 2.6 (1.2) | 0.039 |
| H&Y=1, % (n) | 22.6 (81) | 26 (51) | 18.5 (30) | 0.020 |
| H&Y=2, % (n) | 34.1 (122) | 32.1 (63) | 36.4 (59) | |
| H&Y=3, % (n) | 31.8 (78) | 35.5 (50) | 17.3 (28) | |
| H&Y=4, % (n) | 18.4 (66) | 14.8 (29) | 22.8 (37) | |
| H&Y=5, % (n) | 3.1 (11) | 1.5 (3) | 4.9 (8) | |

The p-values were calculated to compare the responses by gender identity. Mean and standard deviation are reported, unless noted otherwise. Abbreviation: H&Y, Hoehn and Yahr; IQR, interquartile range; SD, standard deviation; n, number.

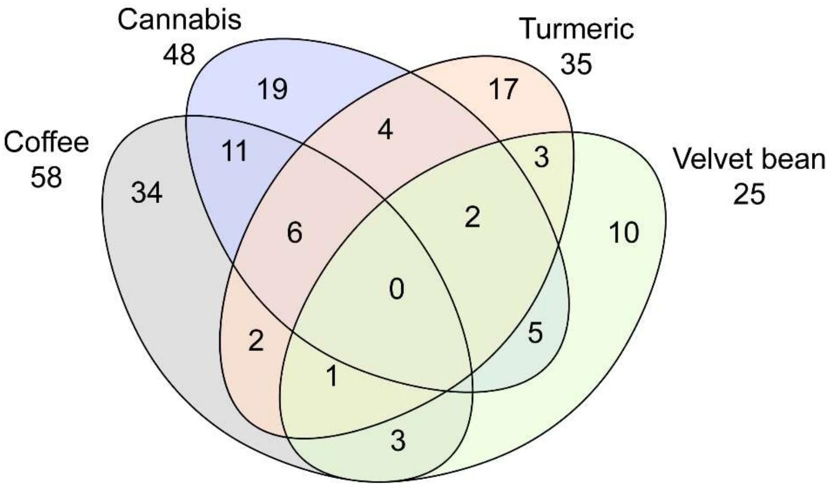


**Supplementary Figure 1. Usage distribution of the top 4 NHP reported by PwP.** Venn diagram illustrating the number of participants who acknowledged using the four most reported herbs listed in Table 1. Each oval represents the use of one product (coffee, cannabis, turmeric and velvet bean). The numbers in these ovals represent the number of participants using the corresponding NHP combination, while the total numbers of participants who use each NHP (either alone or in combination) is provided outside the ovals.
