## Supplementary File 1 for "Plant-based natural products for symptomatic relief of Parkinson’s disease: prevalence, interest, awareness and determinants"

**Questionnaire items:**

i) Have you ever used cannabis products or coffee with the purpose of alleviating symptoms related to Parkinson's Disease? (No / Yes, only cannabis / Yes, only coffee / Yes, both)

ii) Have you ever used any of the following herbal remedies with the purpose of alleviating symptoms related to Parkinson's Disease? You can select more than one option or indicate that you have not used herbal remedies:
   a. Chamomile (e.g *Matricaria recutita, Chamaemelum nobile*)
   b. Curcumin
   c. Guarana
   d. Passion flower (*Passiflora incarnata*)
   e. Resveratrol
   f. Rhodiola (*Rhodiola rosea*)
   g. St. John's Wort (*Hypericum perforatum*)
   h. Turmeric (*Curcuma longa*)
   i. Velvet bean (*Mucuna pruriens*)
   j. Other:
   k. I have not used herbal remedies

iii) Have you ever discussed the possible use of herbal remedies with your neurologist or PD nurse specialist?
   a. Yes
   b. No

iv) Would you be interested in learning more about possible herbal remedies to alleviate certain symptoms related to Parkinson's Disease?
   a. Yes
   b. No

v) Are you aware that some herbal remedies and prescribed Parkinson's Disease medications may work together (or against each other)?
   a. Yes
   b. No
